# Childhood living conditions as predictors of self-rated health status in middle-aged and older adults: Evidence from a machine learning analysis in 27 high-income countries

**DOI:** 10.1101/2025.10.13.25337918

**Authors:** Xu Zong, Karri Silventoinen, Pekka Martikainen, Matti Nelimarkka

**Author notes:** Corresponding author: Xu Zong, Room 308, Unioninkatu 33, Helsinki, Finland, 00170.

## Abstract

**Purpose:** Health inequalities in high-income countries may have roots in childhood. We aimed to develop machine learning (ML) algorithms to assess the impact of childhood living conditions on self-rated health before and during the COVID-19 pandemic.

**Methods:** We analyzed data from 45,570 individuals aged 50+ across 26 European countries and Israel. Information on 39 childhood living conditions and self-rated health was collected. Seven ML algorithms were used to predict health outcomes.

**Results:** All algorithms performed well, with AUC scores ranging from 0.699 to 0.774 and accuracy between 0.621 and 0.724, with CatBoost as the best performer. Self-rated health before age 15 was the most critical predictor. Other key predictors varied by domain such as religion importance, exposure to World War II, physical harm from others and the relationship with the father. We revealed heterogeneity across genders, regions of Europe and levels of COVID-19 lockdown stringency. For instance, mother education was a more prominent predictor of late-life health for females than for males.

**Conclusion:** This study is one of the first to predict health based on childhood conditions and ML. Our results demonstrate the usefulness of ML in identifying early-life conditions that influence health and provide insights for addressing health inequalities.

## 1. Background

In the context of population aging, there is an increasing need to address health inequalities due to their adverse impacts, including heavy healthcare costs, reduced labor productivity, and large welfare costs [1]. There is a strong body of evidence showing that health disparities began in childhood and emphasize the associations between childhood living conditions and late-life inequalities in health [2–6]. Existing studies have generally focused on a few indicators or sets of childhood living conditions, including early socioeconomic position [7], health conditions [8], residential characteristics [9], religious upbringing and exposure to armed conflict or war [10,11].

Moreover, these associations may vary across population subgroups, in this study particularly by genders, regions, and exposure to the COVID-19 pandemic. Numerous studies have identified health inequalities related to genders across Europe, highlighting both the causes and consequences of these disparities [12]. Additionally, the distinct social welfare regimes in different European regions have been shown to be important institutional factors contributing to health inequalities [13]. While the COVID-19 pandemic had a profound global impact, Europe, having one of the oldest populations in the world, experienced a particularly significant impact on population health.

However, it remains unclear whether this external stressor moderated the relationship between early-life living conditions and late-life health.

It is a pressing need to implement early interventions for individuals with adverse childhood living conditions to narrow health inequalities. Effective interventions are contingent upon the accurate evaluation of the importance of each childhood living condition. However, a comprehensive understanding of the relative importance of these predictors is lacking, which may hinder the implementation of effective early interventions, such as identifying individuals at risk of poor health outcomes and offering targeted interventions. A problem in previous studies evaluating the importance of childhood living conditions is that researchers may selectively include childhood variables in their models. Additionally, due to the limitations of statistical methods used in these studies, only a limited number of childhood indicators could be included in the model, since incorporating too many indicators can lead to multicollinearity. Machine learning (ML) offers unique advantages in handling high-dimensional data, enabling the inclusion of a larger number of indicators. This capability not only reduces the risks of subjective variable selection but also reveals the significance of childhood variables that may have been previously overlooked. In this study, we employed ML algorithms to assess the importance of childhood living conditions on health status in adulthood. We studied 39 childhood living conditions across seven domains, including childhood social status, health and healthcare, exposure to wars, abuse, social relationships, residential conditions and cognition in the ML framework to estimate the relative importance of these conditions in predicting late-life health status and examined whether they were more commonly or uniquely observed across genders, regions of Europe and levels of COVID-19 lockdown stringency. Our data was collected from 26 European countries and Israel and include 45,570 participants aged 50 or older.

## 2. Methods

### 2.1 Data

#### 2.1.1 Data Description

We used the Survey of Health, Aging, and Retirement in Europe (SHARE). Comprehensive information about childhood living conditions was collected in the 3rd and 7th wave. Late-life health status was self-reported in the 7th and 9th waves, which were primarily conducted in 2017 and 2021, respectively, corresponding to before and during the COVID-19 pandemic. The ethical reviews for SHARE waves 1 to 4 received approval from the University of Mannheim’s Ethics Committee, and the 4th wave and continuation of SHARE were reviewed and approved by the Ethics Council of the Max Planck Society. More details of the survey design are described elsewhere (Börsch-Supan et al., 2013). Referring to previous studies [14–16], we classified the 26 European countries into four regions with different health and social policy profiles: Northern Europe, Southern Europe, Eastern Europe, and Western Europe. The complete list of countries in each region is presented in Supplementary B. Missing data for childhood living conditions, with an average missing data percentage of 9.45%, along with other variables, were imputed using a random forest algorithm to minimize potential bias caused by imputation. The final analytic sample included 45,570 participants (58.32% women). Flow chart of the analytic sample can be seen in Supplementary F.

#### 2.1.2 Measurements

We used self-rated health to measure health status in late life. Several studies have demonstrated that self-rated health is a valid indicator of health status and is highly correlated with objective health indicators including morbidity and physician assessment [17]. Participants were asked to report their self-rated health status with answers of ‘Excellent’, ‘Very good’, ‘Good’, ‘Fair’, or ‘Poor’. Based on the answers, we created a binary variable, with “Excellent,” “Very good,” and “Good” being categorized as “Good” health (60.95%), while “Fair” and “Poor” were categorized as “Poor” health (39.05%).

To maximize the number of childhood living conditions included in this study, we conducted an in-depth search of the PubMed database to identify published articles using SHARE data to examine the impact of childhood living conditions on late-life health. We selected 39 predictors included in seven domains: childhood social status, childhood health and healthcare, childhood abuse, childhood social relationships, childhood exposure to wars, childhood residential conditions and childhood cognition. A detailed description of these predictors can be found in Supplementary A. Furthermore, we included several covariates in our analysis: age, gender, education, marital status, living in rural or urban areas, living alone, social activities, activities of daily living (ADL), instrumental activities of daily living (IADL), region, and the impact of the COVID-19 pandemic measured by levels of lockdown stringency. We calculated the 30-day average value of the COVID-19 stringency index prior to each participant’s interview using the interview date and daily COVID-19 stringency data from Our World in Data (Mathieu et al., 2020). Supplementary B provides detailed descriptions of these predictors.

### 2.2 Method

#### 2.2.1 Supervised Machine Learning Approach

Supervised ML is an approach to build predictive algorithms based on pre-existing data with dependent and independent variables. It is increasingly used in various disciplines, including sociology, psychology, and biology [18–20]. However, in the field of social health inequalities, studies utilizing ML are still rare. We utilized seven ML classifier algorithms, to evaluate the importance of 39 childhood living conditions in predicting late-life health: Decision trees, random forests, K-Nearest neighbors (KNN), naive bayes, XGBoost, LightGMB and CatBoost. Each algorithm has distinct characteristics [21]. For instance, Decision Trees are simple and interpretable but prone to overfitting; Random forests and XGBoost improve accuracy by aggregating multiple models, with XGBoost offering higher performance and scalability. KNN classifies data based on proximity, while naive bayes uses probabilistic methods assuming feature independence. LightGBM and CatBoost are optimized for speed and handling large or categorical datasets, with LightGBM being faster and CatBoost reducing the need for data preprocessing. Detailed explanations of these algorithms can be found in Supplementary C.

#### 2.2.2. Algorithm Development

Supervised ML algorithms are developed through applying the above-mentioned algorithms to the data, thus finding statistical connections between dependent and independent variables. To avoid overfit and evaluate the generalization ability of an algorithm, data is split into training data, used to develop algorithm, and test data, reserved solely for evaluated its performance. We use a 70%-30% training and testing split. In addition, ML algorithms often do not perform optimally when there are unequal class sizes [22]. This was evident in our data, where only 39.05% of individuals reported having poor health status. Therefore, we used StratifiedKFold to ensure that in each part of our data split, there was a similar proportion of older adults with different health statuses. Additionally, we used grid search methods with 10-fold cross-validation to optimize algorithm hyperparameters.

We compared the predictive performance of these ML algorithms to determine which algorithm provides the optimal predictive performance in identifying the importance of childhood living conditions in predicting health status in late life. We used a set of metrics to evaluate the predictive performance of ML algorithms with the test data set, including accuracy, area under the curve (AUC), sensitivity, specificity, positive predictive values (PPV), and negative predictive values (NPV). While ML algorithms show advanced prediction capabilities, their lack of explanatory ability makes them ‘black boxes’ [19]. To enhance the interpretability of ML algorithms, we used SHapley Additive exPlanations (SHAP), an extension of game theory [20]. SHAP assigns importance values to features in ML algorithms, where positive values mean positive influence on predictive outcomes and negative values mean negative influence, with the magnitude showing the strength of the influence. In this study, we used mean absolute SHAP values to quantify the influence of childhood living conditions on late-life health, providing a robust measure of their importance across the entire dataset.

## 3. Result

### 3.1. Descriptive Analysis

There were 45,570 middle-aged and older individuals (aged 50 years and above) included in both the 7th and 9th waves of SHARE. Before and during the COVID-19 pandemic, disparities in self-rated health were observed in various subgroups, such as age, gender, marital status, education level, living alone and regions. After the onset of COVID-19, the COVID-19 stringency index was associated with self-rated health.

Before COVID-19 (in SHARE wave 7), females (41.00%) exhibited higher rates of poor health compared to males (36.32%) (Supplementary F). The highest percentage of European and Israeli older adults with poor health was observed in the age group of 80 years and older (57.15%), which was significantly higher than in the other age groups. The prevalence of poor health was notably higher among widowed, divorced, and separated individuals (46.64%) compared to their married or partnered counterparts (36.30%). Education also played a crucial role since those with no or primary education exhibited the highest rates of poor health (over 50.00%), whereas those with the second stage of tertiary education had the lowest rates (27.27 %). In the context of welfare regimes, the prevalence ranged from 26.79% among Northern Europeans to 46.78% among Eastern Europeans. The differences between participants having social activities in the last year (28.26%) and participants not having social activities in the last year (45.91%) were significant. During COVID-19 (in SHARE wave 9), these health inequalities still existed. Different from before COVID-19, during the COVID-19 pandemic, individuals having social activities in the last year (47.83%) were more likely to have poor health than those not having social activities in the last year (26.24%), which is opposite to the situation before the pandemic. Additionally, we observed significant disparities in self-rated health among participants who experienced different levels of COVID-19 lockdown stringency: at low stringency level, the percentage of poor self-assessed health was highest (51.78%), but it decreased as the levels increased, then increased again to 44.64% at the highest level. Detailed characteristics of the sample in wave 9 can be found in Supplementary D.

### 3.2 Algorithm Predictive Performance

Supplementary F demonstrate the predictive performance and tuned hyperparameters of the seven ML algorithms evaluating the importance of childhood living conditions in influencing self-rated health in late life. The AUC score was highest for CatBoost (0.774) and lowest for KNN (0.699), both indicating good classification performance. For the accuracy scores, Random forests ranked highest (0.724), followed closely by CatBoost (0.719), with Naive Bayes scoring the lowest (0.621). CatBoost also performed well for other metrics. Overall, CatBoost performed exceptionally well in the classification of good or poor late-life health status. Hence, we employed the CatBoost algorithm to identify the overall and domain-specific importance of conditions and explore the heterogeneity across genders, regions of Europe and COVID-19 lockdown stringency.

### 3.3 Comparison of CatBoost and Logistic Regression

To compare the predictive performances of traditional statistical models and ML algorithm, we conducted a logistic regression analysis using the same predictors and outcomes as the ML algorithm. Supplementary F presents the key predictors of late-life health from childhood living conditions for both the logistic regression and CatBoost algorithm. In logistic regression, exposure to World War I had the largest effect size, but it was not statistically significant. While logistic regression is one of the most commonly used traditional statistical models in previous studies on childhood living conditions and late-life health, CatBoost representing the ML algorithm showed the strongest predictive power in this study. Interestingly, despite their methodological differences, both models highlighted childhood health (health before 15) as critical determinants of health in later years. The logistic regression model produced an accuracy of 0.713 and AUC of 0.755, whereas the CatBoost algorithm achieved accuracy of 0.719 and AUC of 0.774. This indicates that the CatBoost algorithm has a modestly higher predictive performance compared to the logistic regression.

### 3.4 Overall and Domain-specific Importance of Childhood Living Conditions

As shown in Figure 1, among all childhood living conditions, childhood health status (health before age 15) was the strongest predictor of health in late life. Childhood residential conditions (cold/hot running water supply, the number of rooms at age 10) were also critical predictors. We also observed that, for the majority of participants in this study, childhood exposure to war (born during World War II), as well as worse childhood cognition (language and math performance), had a generally negative influence on late-life health. The importance of all childhood and late-life predictors can be seen in Supplementary E.

**Figure 1.**
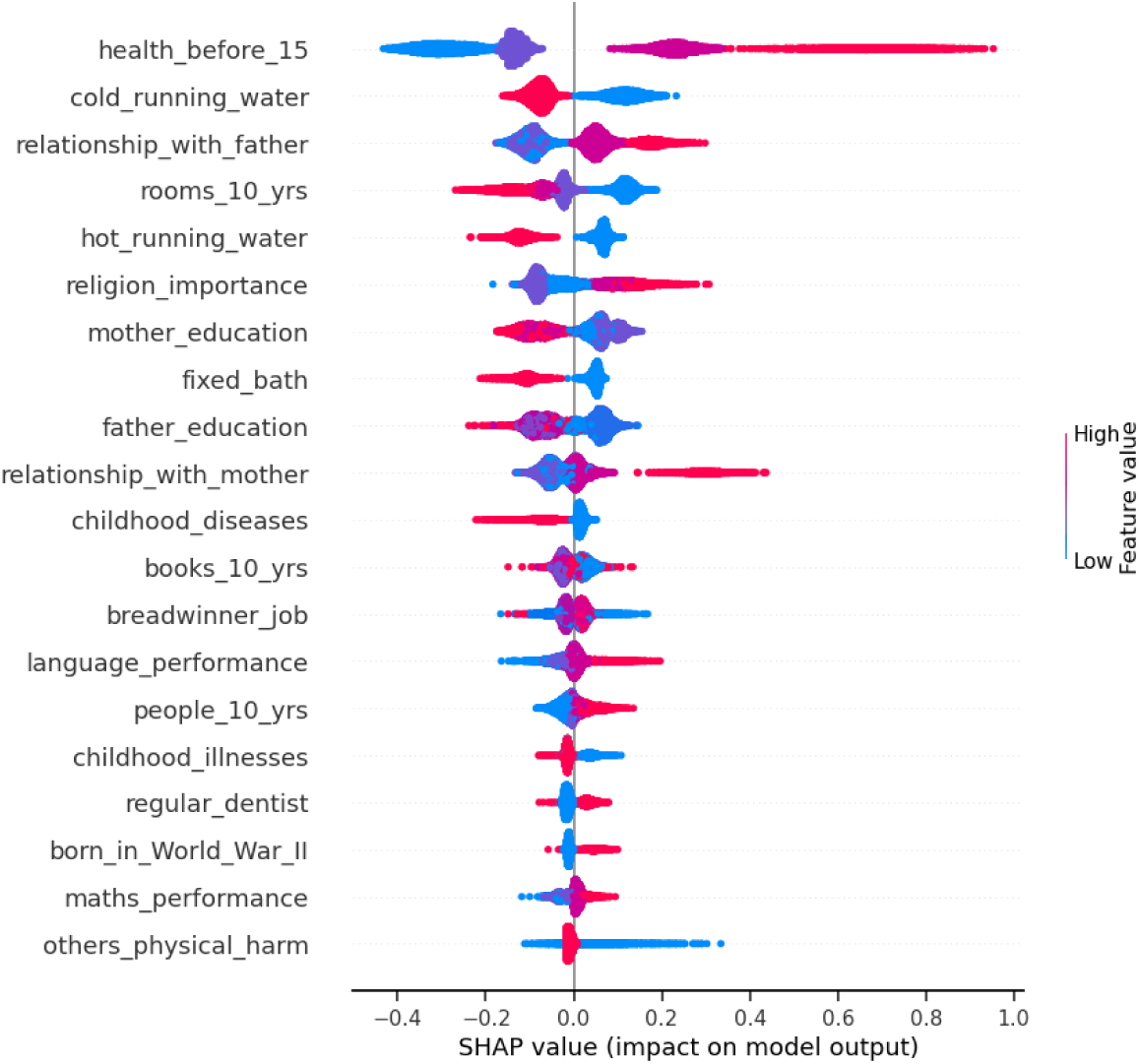
Influence of key childhood living conditions in late-life health Note: SHAP values show the magnitude and direction of these associations with blue points indicating lower values and red points indicating higher values for the conditions on the horizontal axis. They illustrate the importance ranking on the vertical axis, with higher values indicating greater importance. Additionally, the distribution and clustering of SHAP values along the x-axis reflect potential nonlinear relationships between each feature and the outcome. Sharp increases or decreases in SHAP values at specific feature levels suggest threshold or cumulative effects, indicating that the impact on the outcome is not constant but varies significantly across different feature values.

Figure 2 demonstrates the contributions of the seven domains of childhood living conditions to late-life health, with the domain of childhood residential conditions showing the largest impact. In the domain of childhood social status, religion importance (measured by the importance of religion at home when growing up) was identified as the most important predictors of later health, followed by parental education. In the domain of childhood health and healthcare, health before age 15 was identified as the most important predictor, while childhood regular dentist and vaccination had a lasting but smaller influence on late-life health. The domain of childhood exposure to wars showed that exposure to World War II had a persistent impact on health. In the domain of childhood abuse and social relationships, physical harm from others (measured by whether anyone other than the mother or father ever hit the individual) and the relationship with the father identified as the most critical predictors. Regarding residential conditions, availability of cold running water was the strongest predictor, followed the number of books in household at age 10. In the domain of childhood cognition, early language performance was found to be a more predictive predictor of late-life health than early mathematics performance.

**Figure 2.**
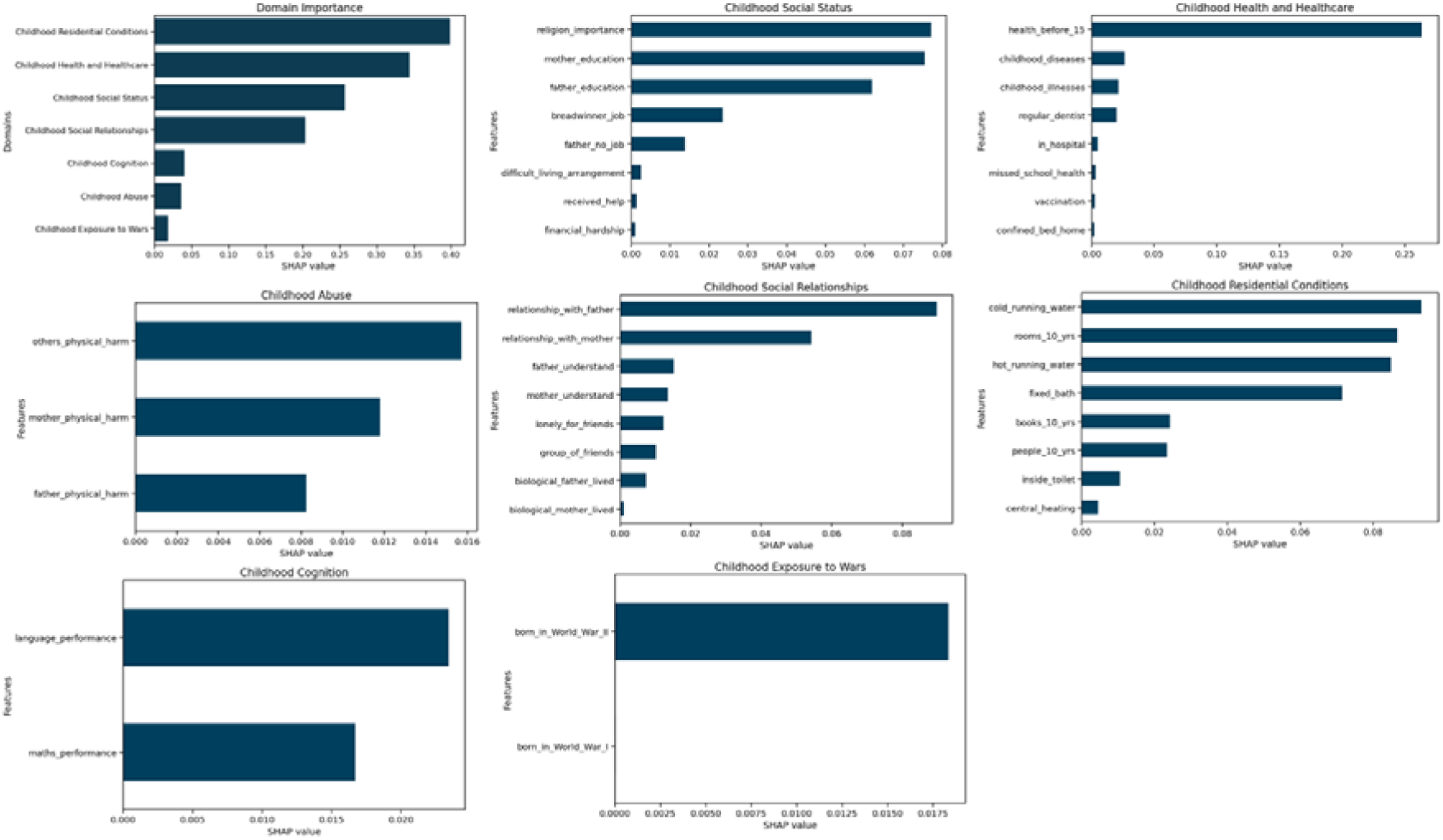
Domain-specific importance of childhood living conditions in late-life health

### 3.5 Heterogeneity of the Importance of Childhood Living Conditions

We analyzed the heterogeneity of childhood living conditions’ impacts on late-life health across genders, regions of Europe, and the COVID-19 lockdown stringency. Figure 3 reveals the shared and distinct predictors across genders. Health before age 15 was the predominant predictor for both males and females. There were notable differences in the influence of other predictors. For females, mother education, math performance, and the number of books at age 10 played more important roles in late-life health compared to males. Figure 4 demonstrates the regional heterogeneity within the regions of Europe of the long-term impact of childhood living conditions. In Northern Europe, parent-child relationships had a large influence on late-life health. In Southern and Eastern Europe, parental education level ranked high as predictors. In Western Europe, early language performance was more key predictor. Figure 5 shows the heterogeneity of various levels of COVID-19 lockdown stringency where higher levels indicate stricter lockdown measures.

**Figure 3.**
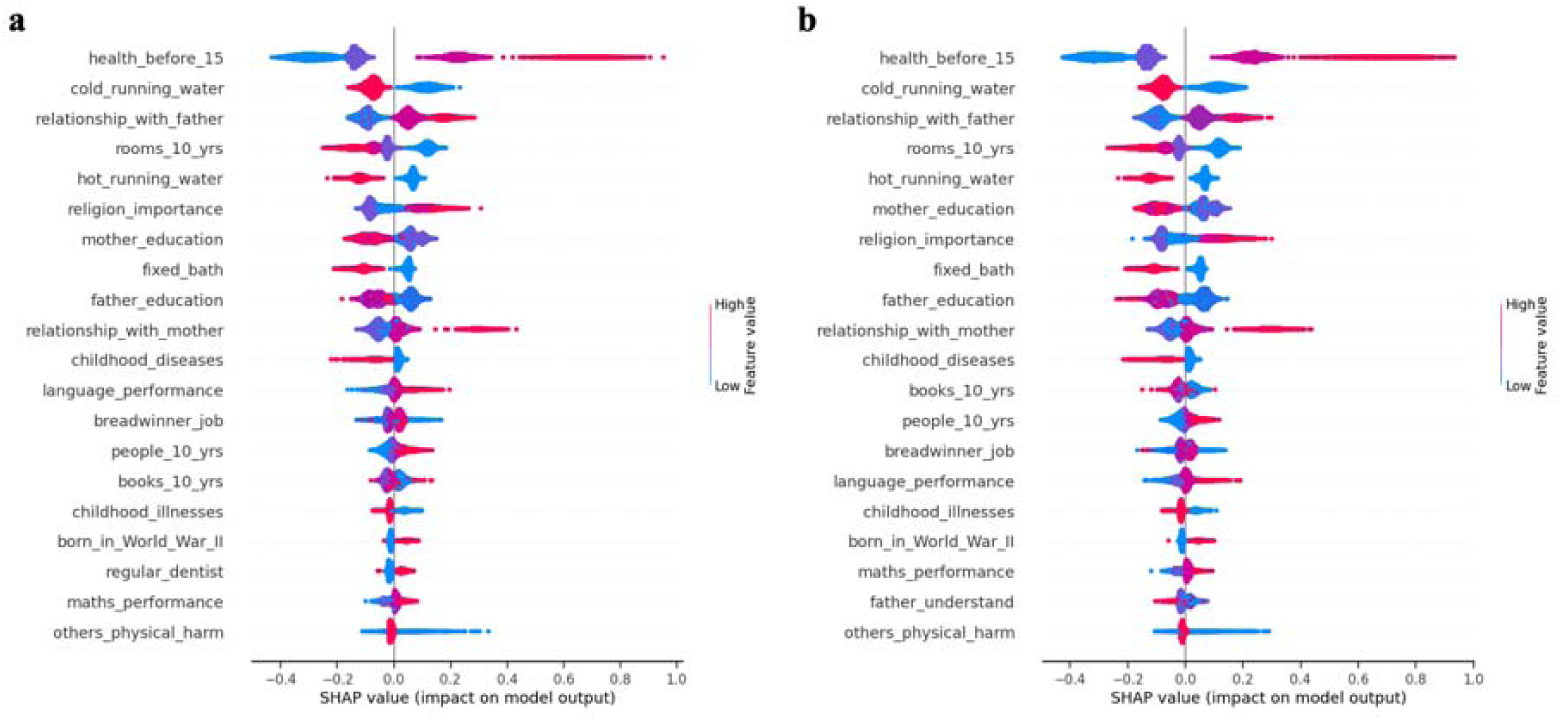
Key predictors for self-rated health in late-life health by genders. Note: ‘a’ represents males, ‘b’ represents females.

**Figure 4.**
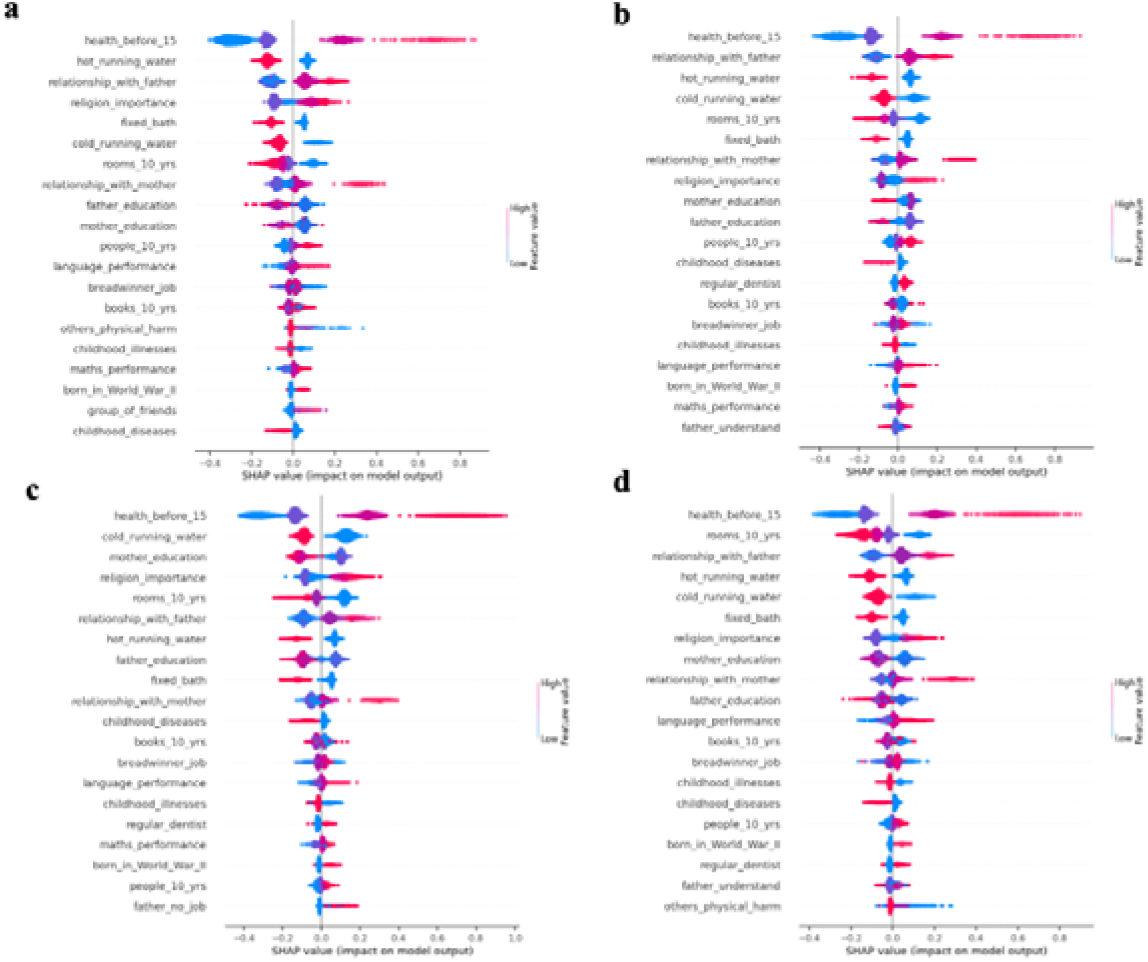
Key predictors for self-rated health in late life by regions Note: ‘a’ represents individuals in Northern Europe, ‘b’ represents individuals in Southern Europe, ‘c’ represents individuals in Eastern Europe, ‘d’ represents individuals in Western Europe.

**Figure 5.**
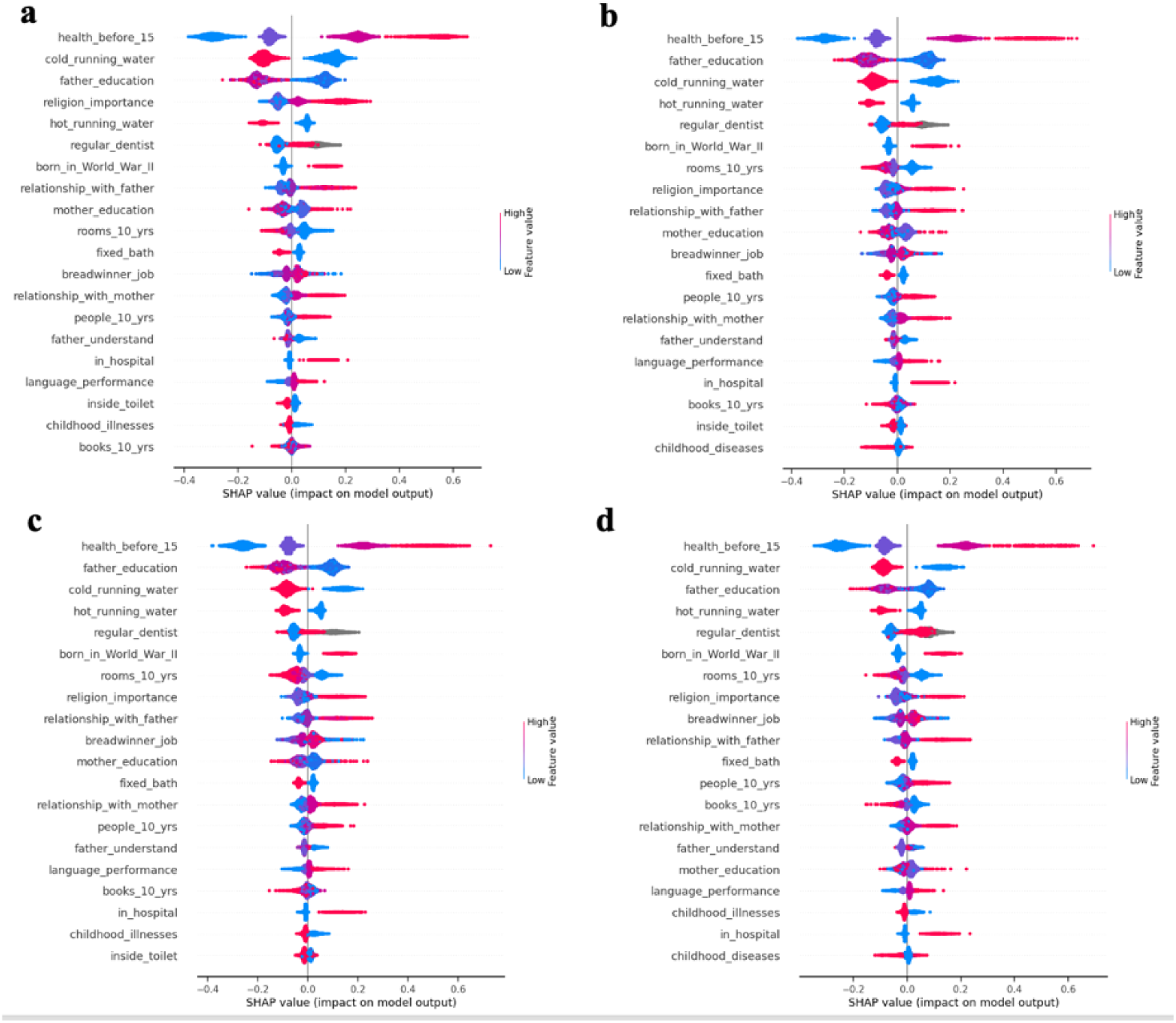
Key predictors for self-rated health in late Life by levels of COVID-19 lockdown stringency. Note: ‘a’ represents individuals experiencing low level of COVID-19 lockdown stringency, ‘b’ represents those experiencing low-to-medium level, ‘c’ represents medium-to-high level, and ‘d’ represents individuals experiencing high level of COVID-19 lockdown stringency.

Compared to the period before the COVID-19 pandemic, the importance ranking of father education and religion importance improved during the low-level lockdown. As the lockdown stringency intensified, early exposure to war became more critical.

## 4. Discussion

To the best of our knowledge, this is the first study to evaluate how a comprehensive set of childhood living conditions contribute to late-life health. By applying multiple ML algorithms to cross-national microdata from 27 countries, we gained further understanding of the relationship between a diverse array of childhood living conditions and late-life health. Firstly, CatBoost outperformed six other ML algorithms, achieving satisfactory predictive performance with an AUC score of 0.774, surpassing those of related studies [23–25]. Furthermore, CatBoost also exceeded the capability of the traditional statistical model (logistic regression) in terms of predictive ability. Secondly, we ranked the leading predictors of all 39 childhood living conditions and identified the most critical predictors within each domain. Religion importance, health before age 15, exposure to World War II, physical harm from others, the relationship with the father, availability of cold running water, and language performance were the top predictors of each domain. Lastly, this study uncovered the heterogeneous influence of childhood living conditions across genders, regions of Europe, and COVID-19 lockdown stringency.

Our findings emphasized the lasting influence of various childhood living conditions, aligning with earlier studies on the health impacts of early-life conditions [26–29]. This supports the importance of incorporating life-course perspectives in the study of health inequalities and policy-making. By including as many childhood living conditions as possible, we identified some critical predictors that have not been given sufficient attention in previous studies including a limited number of childhood living conditions. These predictors included early cognition (language and math performance), childhood residential conditions (cold/hot water supply, the number of rooms at age 10), and religion importance. Interestingly, our findings indicate that poor childhood cognition had generally negative associations with late-life health in Europe, which aligns with a study reporting that childhood cognition (academic performance) was negatively associated with adverse adult health [30]. Additionally, our findings demonstrate that poor residential conditions are associated with adverse health outcomes, consistent with studies linking housing conditions to inequalities in mortality, cancer, and mental health [31,32]. The potential mechanisms behind these associations warrant further studies.

The COVID-19 pandemic profoundly affected human health, especially among the older population [33,34]. This is the first study that explored the influence of a broad range of childhood living conditions on the health of middle-aged and older adults in Europe during the pandemic. We identified the varying impact of these conditions across levels of lockdown stringency, demonstrating that early-life circumstances continued to play a crucial role in health during the pandemic. This aligns with a UK study that found close associations between childhood circumstances and COVID-19-related mortality [35]. Our findings provide more comprehensive and precise evidence for policy-making to improve the health and well-being of older adults by screening individuals who experienced various adverse childhood living conditions in future public health crises similar to the COVID-19 pandemic. Specifically, our analysis revealed that the importance of certain childhood conditions, such as father education and religion importance, increased during low-level lockdowns, while factors like early exposure to war became more critical under stricter lockdown measures. In low-level lockdowns, where social activities were moderately restricted but still ongoing, well-educated fathers may have provided their families with greater economic and social resources, helping them better manage stress and alleviate mental health burdens. Additionally, religion might offer emotional comfort and spiritual support. However, under a stricter lockdown, the restrictions on daily life, scarcity of supplies, and lack of freedom might evoke memories of wartime experiences for those affected, triggering intense psychological stress responses. This reactivation of trauma might exacerbate mental strain, leading to poorer health outcomes.

Our study has advantages in five aspects. First, we employed multiple ML algorithms and selected the optimal ML algorithm to provide the relative importance of every childhood living condition in late life, which is challenging to analyze using traditional statistical models due to methodological limitations. Second, creating 39 childhood living conditions across seven domains made this study include a more comprehensive set of indicators of childhood living conditions compared to existing related studies using SHARE data. Third, we compared the predictive ability between the traditional statistical model (logistic regression) and ML algorithm (CatBoost) and the common predictors in the two models. Fourth, we further revealed the positive or negative health impact of childhood living conditions and the impact extent by leveraging SHAP. Fifth, we considered the heterogeneous impacts of childhood living conditions across different genders, regions of Europe and COVID-19 lockdown stringency.

There are several limitations in this study. First, the measurements of childhood living conditions and late-life health were self-reported respectively, and hence may lead to recall and reporting bias of measures [36]. Second, survival bias my affect out results since individuals who experienced serious childhood adversities may have died prematurely [37]. Third, while existing studies have shown that childhood adversities often cluster and have long-term systematic influences [38], this study focused on the importance evaluation of single childhood living conditions across seven domains, thus making no claims about causal pathways. Future research should thus further explore the long-term influence of childhood living conditions by adopting more objective measurements and considerations of cumulative effects.

## 5. Conclusion

This study comprehensively analyzed the overall and specific contributions of 39 childhood living conditions to health among older European and Israeli populations. It confirms the close associations between childhood living conditions and late-life health, while also highlighting the heterogeneous impacts of these conditions across genders, regions of Europe, and levels of COVID-19 lockdown stringency. For example, health before age 15 was a predominant predictor for both genders, but mother education was more significant for females. Regionally, parent-child relationships were more influential in Northern Europe, while maternal education and the breadwinner’s job had greater effects in Southern and Eastern Europe. The stringency of COVID-19 lockdowns also revealed differences: under stricter lockdowns, early exposure to war became a more critical factor, while the relevance of father’s education decreased. Our findings emphasize the pressing need for targeted strategies to mitigate childhood adversity to improve late-life health from the life course perspective, taking into account these heterogeneities.

## Declarations

### Ethics approval and consent to participate

The ethical reviews for SHARE waves 1 to 4 received approval from the University of Mannheim’s Ethics Committee, and the 4th wave and continuation of SHARE were reviewed and approved by the Ethics Council of the Max Planck Society.

## Consent for publication

Not applicable.

## Availability of data and materials

The dataset analyzed in this study is publicly available at the following website: https://share-eric.eu/.

## Authors’ contributions

XZ’s contributions include: conceptualisation, data curation, formal analysis, funding acquisition, investigation, methodology, project administration, resources, software, validation, visualisation, writing - original draft, and writing - review & editing.

KS’s contributions include: project administration, writing - original draft, and writing - review & editing.

PM’s contributions include: funding acquisition, writing - original draft, and writing - review & editing.

MN’s contributions include: methodology, software, writing - original draft, and writing - review & editing.

## Data Availability

The SHARE data used in this study can be assessed at https://share-eric.eu/.

## Acknowledgements

We would like to express our sincere gratitude to the Transnational Access Visits (TAV) as part of the COORDINATE project (Grant Agreement Number: 101008589) and European Centre for Social Welfare Policy and Research for their invaluable support in providing training on data related to this research. This training significantly enhanced our understanding of the dataset used in this study.

This paper uses data from SHARE Waves 3, 7 and 9 (DOIs: 10.6103/SHARE.w1.900, 10.6103/SHARE.w3.900, 10.6103/SHARE.w7.900, 10.6103/SHARE.w9.900) see Börsch-Supan et al. (2013) for methodological details. The SHARE data collection has been funded by the European Commission, DG RTD through FP5 (QLK6-CT-2001-00360), FP6 (SHARE-I3: RII-CT-2006-062193, COMPARE: CIT5-CT-2005-028857, SHARELIFE: CIT4-CT-2006-028812), FP7 (SHARE-PREP: GA N°211909, SHARE-LEAP: GA N°227822, SHARE M4: GA N°261982, DASISH: GA N°283646) and Horizon 2020 (SHARE-DEV3: GA N°676536, SHARE-COHESION: GA N°870628, SERISS: GA N°654221, SSHOC: GA N°823782, SHARE-COVID19: GA N°101015924) and by DG Employment, Social Affairs & Inclusion through VS 2015/0195, VS 2016/0135, VS 2018/0285, VS 2019/0332, VS 2020/0313, SHARE-EUCOV: GA N°101052589 and EUCOVII: GA N°101102412. Additional funding from the German Federal Ministry of Education and Research (01UW1301, 01UW1801, 01UW2202), the Max Planck Society for the Advancement of Science, the U.S. National Institute on Aging (U01_AG09740-13S2, P01_AG005842, P01_AG08291, P30_AG12815, R21_AG025169, Y1-AG-4553-01, IAG_BSR06-11, OGHA_04-064, BSR12-04, R01_AG052527-02, R01_AG056329-02, R01_AG063944, HHSN271201300071C, RAG052527A) and from various national funding sources is gratefully acknowledged (see www.share-eric.eu).

