## Supplementary A-Supplementary F for "Childhood living conditions as predictors of self-rated health status in middle-aged and older adults: Evidence from a machine learning analysis in 27 high-income countries"

Detailed description of these predictors of childhood living conditions.

| Domains | Predictors | Meaning of predictors |
| --- | --- | --- |
| Childhood social status | mother_education | The education level of respondent’s biological mother  0. None  1. Primary education  2. Lower secondary education  3. Upper secondary education  4. Post-secondary non tertiary education  5. First stage of tertiary education  6. Second stage of tertiary education |
|  | father_education | The education level of respondent’s biological father  0. None  1. Primary education  2. Lower secondary education  3. Upper secondary education  4. Post-secondary non tertiary education  5. First stage of tertiary education  6. Second stage of tertiary education |
|  | breadwinner_job | The occupation of main breadwinner when respondent was 10 years  1. Legislator, senior official or manage  2. Professional  3. Technician or associate professional  4. Clerk  5. Service, shop or market sales worker  6. Skilled agricultural or fishery worke  7. Craft or related trades worker  8. Plant/machine operator or assembler  9. Elementary occupation  10. Armed forces  11. Spontaneous only: there was no main |
|  | financial_hardship | Whether respondent experienced financial hardship before age 16  0. No  1. Yes |
|  | received_help | Before age 16, whether there was a time when respondent or the respondent’s family received help from relatives because of financial difficulties  0.No  1.Yes |
|  | difficult_living_arrangement | Whether respondent experienced difficult living arrangement before age 16  0. No  1. Yes |
|  | father_no_job | Before age 16, whether there was a time of several months or more when the respondent’s father had no job  1.Yes  2.No  6.Father never worked or always disabled  7.Never lived with father or father was not alive |
|  | religion_importance | The importance of religion at home when respondent grew up  1. Very important  2. Somewhat important  3. Not very important  4. Not at all important |
| Childhood health and healthcare | health_before_15 | Respondent’s self-reported health status during childhood  1. Excellent  2. Very good  3. Good  4. Fair  5. Poor  6. Health varied a great deal |
|  | missed_school_health | Whether respondent missed school for 1+ months due to health before age 16  0. No  1. Yes |
|  | confined_bed_home | Whether respondent was confined to bed or home for 1 month or longer  0. No  1. Yes |
|  | childhood_diseases | Whether respondent had any of the diseases before age 16 (Infectious disease, Polio, Asthma, Respiratory problems other than asthma, Allergies, Severe diarrhoea, Meningitis/encephalitis, Chronic ear problems, Speech impairment, Difficulty seeing even with eyeglasses)  0. No  1. Yes |
|  | childhood_illnesses | Whether respondent had any of the illness before age 16 (Severe headaches or migraines, Epilepsy, fits or seizures, Emotional, nervous, or psychiatric problem, Broken bones, fractures, Appendicitis, Childhood diabetes or high blood sugar, Heart trouble, Leukaemia or lymphoma, Cancer or malignant tumour (excluding minor skin cancers), Rickets, osteomalacia, rachitis)  0. No  1. Yes |
|  | in_hospital | During childhood, because of a health condition, whether respondent ever was in hospital for one month or more  5. No  1. Yes |
|  | vaccination | Whether respondent had vaccination in childhood  0. No  1. Yes |
|  | regular_dentist | Whether respondent started going regularly to the dentist during your childhood (that is, before you turned 16)  5. No  1. Yes |
| Childhood exposure to wars | born_in_World_War_I | Whether respondent was born between 1914 and 1918  0. No  1.Yes |
|  | born_in_World_War_II | Whether respondent was born between 1939 and 1945  0. No  1. Yes |
| Childhood abuse | mother_physical_harm | When you were growing up, did your female guardian ever hit you?  1. Often  2. Sometimes  3. Rarely  4. Never |
|  | father_physical_harm | When you were growing up, did your male guardian ever hit you?  1. Often  2. Sometimes  3. Rarely  4. Never |
|  | others_physical_harm | When you were growing up, how often did your brother or sister ever hit you?  1. Often  2. Sometimes  3. Rarely  4. Never |
| Childhood social relationships | lonely_for_friends | How often respondent felt lonely for friends in childhood  1. Often  2. Sometimes  3. Rarely  4. Nerve |
|  | group_of_friends | Between ages 6-16, how often respondent had a group of friends that felt comfortable spending time with  1. Often  2. Sometimes  3. Rarely  4. Nerve |
|  | relationship_with_mother | How would you rate your relationship with your female guardian when you were growing up?  1. Excellent  2.Very good  3.Good  4.Fair  5.Poor |
|  | mother_understand | How much did your mother understand your problems and worries  1.A lot  2.Some  3.A little  4.Not at all |
|  | relationship_with_father | How would you rate your relationship with your male guardian when you were growing up?  1. Excellent  2.Very good  3.Good  4.Fair  5.Poor |
|  | father_understand | How much did your father understand your problems and worries  1.A lot  2.Some  3.A little  4.Not at all |
|  | biological_mother_lived | Whether respondent’s mother lived when respondent was 10 years  0. No  1. Yes |
|  | biological_father_lived | Whether respondent’s father lived with when respondent was 10 years  0. No  1. Yes |
| Childhood residential conditions | rooms_10_yrs | This predictor represents the number of rooms in the accommodation where the respondent lived when they were 10 years old. The values are divided into quartiles.  1.First quartile  2.Second quartile  3.Third quartile  4.Fourth quartile |
|  | people_10_yrs | This predictor represents the number of people in the household where the respondent lived when they were 10 years old. The values are divided into quartiles.  1.First quartile  2.Second quartile  3.Third quartile  4.Fourth quartile |
|  | books_10_yrs | This predictor represents the number of books in the household where the respondent lived when they were 10 years old. The values are divided into quartiles.  1.First quartile  2.Second quartile  3.Third quartile  4.Fourth quartile |
|  | cold_running_water | Whether there had cold running water supply in accommodation when respondent was 10 years  0. No  1. Yes |
|  | hot_running_water | Whether there had hot running water supply in accommodation when respondent was 10 years  0. No  1. Yes |
|  | fixed_bath | Whether there had fixed bath in accommodation when respondent was 10 years  0. No  1. Yes |
|  | inside_toilet | Whether there had inside toilet in accommodation when respondent was 10 years  0. No  1. Yes |
|  | central_heating | Whether there had central heating in accommodation when respondent was 10 years  0. No  1. Yes |
| Childhood cognition | maths_performance | Respondent’s relative performance of mathematics when 10  1. Much better  2. Better  3. About the same  4. Worse  5. Much worse |
|  | language_performance | Respondent’s relative performance of language when 10  1. Much better  2. Better  3. About the same  4. Worse  5. Much worse |

### Supplementary B

Detailed description of late-life sociodemographic predictors

| Domains | Predictors | Meaning of predictors |
| --- | --- | --- |
|  | gender | Respondent’s gender  1. Man  0. Woman |
|  | age | Respondent’s age at interview  1.50-59 years old  2.60-69 years old  3.70-79 years old  4.80+ years old |
|  | marriage | Respondent’s marital status  1. Separated, divorced, widowed, never married  0. Married or registered partnership |
|  | education | Respondent’s education level  0. None  1. Primary education  2. Lower secondary education  3. Upper secondary education  4. Post-secondary non tertiary education  5. First stage of tertiary education  6. Second stage of tertiary education |
|  | living in rural or urban areas | Whether respondent lives in rural or urban area  0. Urban  1. Rural |
|  | living alone | Whether respondent lives alone  0. No  1. Yes |
|  | region | Region where the respondent lives  1. Northern European countries (Denmark, Sweden, Finland)  2. Southern European countries (Spain, Italy, Greece, Cyprus, Malta, Portugal)  3. Eastern European countries (Slovenia, Estonia, Lithuania, Bulgaria, Latvia, Romania, Poland, Hungary, Slovakia, Czech Republic, Croatia)  4. Western European countries (France, Belgium, Ireland, Luxembourg, Austria, Germany, Switzerland)  5. Israel |
|  | social | Whether the respondent has any yearly social activities    0. No  1. Yes |
|  | ADL | Number of limitations with activities of daily living |
|  | IADL | Number of limitations with instrumental activities of daily living |

Levels of COVID-19 lockdown stringency was calculated as the mean score of nine metrics: school closures; workplace closures; cancellation of public events; restrictions on public gatherings; closures of public transport; stay-at-home requirements; public information campaigns; restrictions on internal movements; and international travel controls (Mathieu et al., 2020). The higher the index is, the stricter the response is.

Supplementary C

Explanations of the ML algorithms

**Decision trees**

Decision trees (DT) is a ML algorithm that is used for classification and regression. DT will construct tree-like models, each node of the tree stands for a decision, and the data is split by the decision. Finally, the splits will come to the terminal nodes of the tree, also called ‘leaves’, which stands for the final classification outcomes. DTs are easy to interpret because tress can be visualized, but they tend to be overfitting when trees are too complex.

**Random forests**

Random forests (RF) is a kind of ensemble learning algorithm that is an extension of the decision trees. RF develop many decision tress and create the modes of classification of the single trees to make prediction. RF calculate the averages of multiple tress to reduce the variance, which decrease the risk of being overfitting. RF is good at dealing with numerical and categorical data, handling large data and missing data, and usually perform better than single decision trees, while it lacks explainability and cost relatively much computational resources.

**K-Nearest neighbors**

K-Nearest neighbors (KNN) is a non-parametric ML algorithm that usually work for classification, regression, and imputing missing data. The basic idea is that, when we need to classify a new data point, KNN will calculate the distance between this data point with other data points among the training dataset, then we select the nearest data points also called neighbors, finally we use majority vote method to decide the classification of the new data point. KNN is adaptive to classification and regression tasks but sensitive to noisy data and computationally intensive.

**Naïve bayes**

Naïve bayes (NB) is ML algorithm predominantly used for classification tasks. It is based on Bayes’ theorem, and fundamental idea is behind it is to make classification by calculating the conditional probability of each class based on the features and then making selection of the class with the highest probability. NB is easy and efficient to perform, and good at handing high-dimensional data and noisy data. It has some limits, such as independence assumption that is often hard to satisfy, and sensitive to deal with class imbalanced data.

**Light gradient boosting machine**

Light gradient boosting machine (LightGBM) is developed by Microsoft to perform classification and regression tasks, especially suitable for large datasets. It is grounded in the principles of gradient boosting. It has several advantages such as handling with large datasets, training models efficiently, allowing parallel and distributed learning. It is prone to be overfitting, sensitive to noisy data, challenging to turn parameters, and lack interpretability,

**eXtreme Gradient Boosting**

eXtreme Gradient Boosting (XGBoost) is a powerful ML algorithm due to exceptional performance and efficiency. It is based on the gradient boosting framework. It is known for often outperforming other ML models in prediction, especially for large datasets and complex tasks. Besides, it is good at dealing with missing data. For XGBoost, there are some limits, including risk of overfitting, time-consuming to train models and tune hyperparameters, and difficult to interpret.

**Categorical boosting**

Categorical boosting (CatBoost) is a ML algorithm based on gradient boosting, originally developed by Yandex. It is notable that CatBoost can deal with categorical features directly and efficiently and hence result in less data preprocessing. Several advantages made CatBoost perform well in prediction, such as handling categorical features, ordered boosting, and efficient training. It may lead to the complexity of hyperparameter tuning.

Supplementary D

Demographic Characteristics of Participants of wave 9 in this study

| Characteristics | having good health | | Having poor health | | Total | | p-value |
| --- | --- | --- | --- | --- | --- | --- | --- |
|  | n | % | n | % | n | % |  |
| Total |  |  |  |  |  |  |  |
| **Gender** |  |  |  |  |  |  | <0.001 |
| Male | 11,419 | 60.12 | 7,576 | 39.88 | 18,995 | 41.68 |  |
| Female | 15,276 | 57.48 | 11,299 | 42.52 | 26,575 | 58.32 |  |
| **Age** |  |  |  |  |  |  | <0.001 |
| **50-59** | 2,481 | 71.89 | 970 | 28.11 | 3,451 | 7.57 |  |
| **60-69** | 11,355 | 68.66 | 5,182 | 31.34 | 16,537 | 36.29 |  |
| **79-79** | 9,230 | 56.01 | 7,250 | 43.99 | 16,480 | 36.10 |  |
| **80+** | 3,629 | 39.87 | 5,473 | 60.13 | 9,102 | 19.97 |  |
| **Marriage** |  |  |  |  |  |  | <0.001 |
| Married or partnered | 18,583 | 62.05 | 11,366 | 37.95 | 29,949 | 65.72 |  |
| Separated, divorced, widowed, never married | 8,112 | 51.93 | 7,509 | 48.07 | 15,621 | 34.28 |  |
| **Education** |  |  |  |  |  |  | <0.001 |
| None | 700 | 43.21 | 920 | 56.79 | 1,620 | 3.54 |  |
| Primary education | 2,692 | 43,42 | 3,508 | 56.58 | 6,200 | 13.58 |  |
| Lower Secondary education | 4,135 | 52.80 | 3,697 | 47.20 | 7,832 | 17.17 |  |
| Upper secondary education | 10,513 | 61.93 | 6,463 | 38.07 | 16,976 | 37.29 |  |
| Post-secondary non tertiary education | 1,358 | 56.47 | 1,047 | 43.53 | 2,405 | 5.28 |  |
| First stage of tertiary education | 7,063 | 69.42 | 3,111 | 30.58 | 10,174 | 22.35 |  |
| Second stage of tertiary education | 234 | 64.46 | 129 | 35.54 | 363 | 0.80 |  |
| **Living in rural or urban areas** |  |  |  |  |  |  | <0.001 |
| Urban | 18,033 | 59.72 | 12,162 | 40.28 | 30,195 | 66.26 |  |
| Rural | 8,662 | 56.34 | 6,713 | 43.66 | 15,375 | 33.74 |  |
| **Social** |  |  |  |  |  |  | <0.001 |
| No | 9,985 | 73.76 | 3,553 | 26.24 | 13,538 | 29.71 |  |
| Yes | 16,710 | 52.17 | 15,322 | 47.83 | 32,032 | 70.29 |  |
| **Living alone** |  |  |  |  |  |  | <0.001 |
| No | 20,205 | 61.09 | 12,871 | 38.91 | 33,076 | 72.58 |  |
| Yes | 6,790 | 51.94 | 6,004 | 48.06 | 12,494 | 27.42 |  |
| **Region** |  |  |  |  |  |  | <0.001 |
| Northern Europe | 3,521 | 69.50 | 1,545 | 30.50 | 5,066 | 11.12 |  |
| Southern Europe | 4,865 | 54.33 | 4,090 | 45.67 | 8,955 | 19.65 |  |
| Eastern Europe | 10.110 | 52.74 | 9,059 | 47.26 | 19,169 | 42.06 |  |
| Western Europe | 7,941 | 66.70 | 3,964 | 33.30 | 11,905 | 26.16 |  |
| Israel | 258 | 54.32 | 217 | 45.68 | 475 | 1.04 |  |
| **COVID-19 lockdown stringency** |  |  |  |  |  |  | <0.001 |
| Low level | 5,544 | 48.22 | 5,953 | 51.78 | 11,497 | 25.23 |  |
| Low-to-medium level | 6,848 | 63.49 | 3,938 | 36.51 | 10,786 | 23.67 |  |
| Medium-to-high level | 8,403 | 66.53 | 4,227 | 33.47 | 12,630 | 27.72 |  |
| High level | 5,900 | 55.36 | 4,757 | 44.64 | 10,657 | 23.39 |  |

Supplementary E

The importance of all childhood living conditions and late-life sociodemographic features in SHARE wave 7.

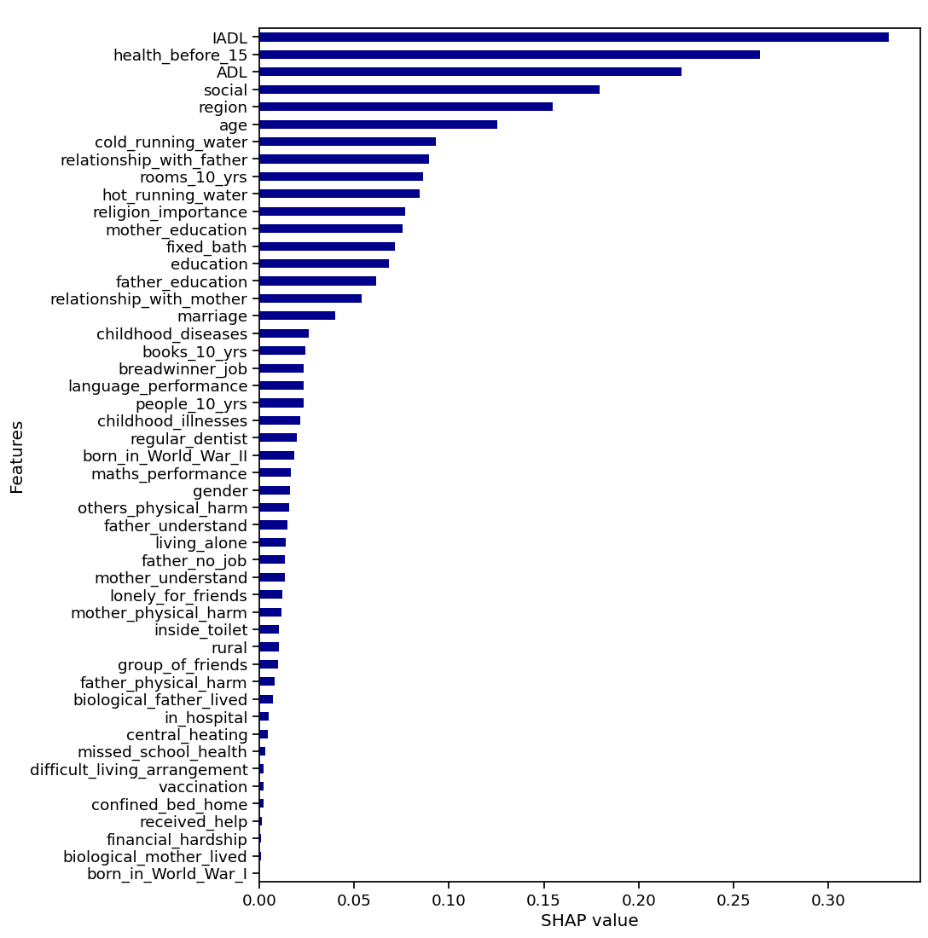

Supplementary F

Table. Demographic Characteristics of Participants of wave 7 in this study

| **Characteristics** | **having good health** | | **Having poor health** | | **Total** | | **p-value** |
| --- | --- | --- | --- | --- | --- | --- | --- |
|  | n | % | n | % | n | % |  |
| Total |  |  |  |  |  |  |  |
| **Gender** |  |  |  |  |  |  | <0.001 |
| Male | 12,096 | 63.68 | 6,899 | 36.32 | 18,995 | 41.68 |  |
| Female | 15,680 | 59.00 | 10,895 | 41.00 | 26,575 | 58.32 |  |
| **Age** |  |  |  |  |  |  | <0.001 |
| 50-59 | 7,201 | 70.08 | 3,075 | 29.92 | 10,276 | 22.55 |  |
| 60-69 | 11,904 | 64.68 | 6,500 | 35.32 | 18,404 | 40.39 |  |
| 79-79 | 6,898 | 54.09 | 5,854 | 45.91 | 12,752 | 27.98 |  |
| 80+ | 1,773 | 42.85 | 2,365 | 57.15 | 4,138 | 9.08 |  |
| **Marriage** |  |  |  |  |  |  | <0.001 |
| Married or partnered | 21,317 | 63.70 | 12,149 | 36.30 | 33.466 | 73.44 |  |
| Separated, divorced, widowed, never married | 6,459 | 53.36 | 5,645 | 46.64 | 12,104 | 26.56 |  |
| **Education** |  |  |  |  |  |  | <0.001 |
| None | 765 | 47.22 | 855 | 52.78 | 1,620 | 3.55 |  |
| Primary education | 3,019 | 48.69 | 3,181 | 51.31 | 6,200 | 13.61 |  |
| Lower Secondary education | 4,245 | 54.20 | 3,587 | 45.80 | 7,832 | 17.19 |  |
| Upper secondary education | 10,790 | 63.56 | 6,186 | 36.44 | 16,976 | 37.25 |  |
| Post-secondary non tertiary education | 1,367 | 56.84 | 1,038 | 43.16 | 2,405 | 5.28 |  |
| First stage of tertiary education | 7,326 | 72.01 | 2,848 | 27.99 | 10,174 | 22.33 |  |
| Second stage of tertiary education | 264 | 72.73 | 99 | 27.27 | 363 | 0.80 |  |
| **Living in rural or urban areas** |  |  |  |  |  |  | <0.001 |
| Urban | 17,420 | 61.91 | 10,718 | 38.09 | 28,138 | 61.75 |  |
| Rural | 10,356 | 59.41 | 7,076 | 40.59 | 17,432 | 38.25 |  |
| **Social** |  |  |  |  |  |  | <0.001 |
| No | 15,065 | 54.09 | 12,788 | 45.91 | 27,853 | 61.12 |  |
| Yes | 12,711 | 71.74 | 5,006 | 28.26 | 17,717 | 38.88 |  |
| **Living alone** |  |  |  |  |  |  | <0.001 |
| No | 22,419 | 62.85 | 13,254 | 37.15 | 35,673 | 78.28 |  |
| Yes | 5,357 | 54.13 | 4,540 | 45.87 | 9,897 | 21.72 |  |
| **Region** |  |  |  |  |  |  | <0.001 |
| Northern Europe | 3,709 | 73.21 | 1,357 | 26.79 | 5,066 | 11.12 |  |
| Southern Europe | 5,303 | 59.22 | 3,652 | 40.78 | 8,955 | 19.65 |  |
| Eastern Europe | 10.202 | 53.22 | 8,967 | 46.78 | 19,169 | 42.06 |  |
| Western Europe | 8,283 | 69.58 | 3,622 | 30.42 | 11,905 | 26.12 |  |
| Israel | 279 | 58.74 | 196 | 41.26 | 475 | 1.04 |  |

Table. Algorithm predictive performance on the test data

| **Algorithm** | **AUC** | **Accuracy** | **Sensitivity** | **Specificity** | **PPV** | **NPV** |
| --- | --- | --- | --- | --- | --- | --- |
| CatBoost | 0.774 | 0.719 | 0.649 | 0.764 | 0.636 | 0.775 |
| Decision trees | 0.741 | 0.707 | 0.424 | 0.886 | 0.702 | 0.708 |
| Random forests | 0.772 | 0.724 | 0.571 | 0.821 | 0.669 | 0.751 |
| KNN | 0.699 | 0.678 | 0.386 | 0.863 | 0.642 | 0.689 |
| Naive bayes | 0.726 | 0.621 | 0.781 | 0.520 | 0.508 | 0.789 |
| XGBoost | 0.773 | 0.717 | 0.652 | 0.759 | 0.631 | 0.775 |
| LightBGM | 0.774 | 0.717 | 0.652 | 0.758 | 0.631 | 0.774 |

Table. Tuned hyperparameters

| **Algorithm** | **Best Parameters** | **Parameter range** |
| --- | --- | --- |
| CatBoost | 'depth': 6,  'iterations': 300,  'learning_rate': 0.05} | 'depth': [4, 6, 8],  'iterations': [100, 200, 300],  'learning_rate': [0.01, 0.05, 0.1] |
| Decision trees | 'max_depth': 7,  'min_samples_leaf': 4,  'min_samples_split': 10 | 'max_depth': [3, 5, 7, 10, None],  'min_samples_leaf': [1, 2, 4],  'min_samples_split': [2, 5, 10] |
| Random forests | 'max_depth': 30,  'min_samples_split': 10,  'n_estimators': 200 | 'max_depth': [10, 20, 30],  'min_samples_split': [2, 5, 10],  'n_estimators': [50, 100, 200] |
| KNN | 'metric': 'manhattan',  'n_neighbors': 11,  'weights': 'distance' | 'metric': ['euclidean', 'manhattan'],  'n_neighbors': [3, 5, 7, 9, 11],  'weights': ['uniform', 'distance'] |
| Naive bayes | / | / |
| XGBoost | 'learning_rate': 0.1,  'max_depth': 3,  'n_estimators': 200} | 'learning_rate': [0.01, 0.05, 0.1],  'max_depth': [3, 5, 7],  'n_estimators': [50, 100, 200] |
| LightGBM | 'learning_rate': 0.05,  'max_depth': 6,  'n_estimators': 200} | 'learning_rate': [0.01, 0.05, 0.1]  'max_depth': [4, 6, 8],  'n_estimators': [100, 200, 300] |

Note: In this study, we used the GaussianNB classifier for the naive bayes algorithm, GaussianNB does not require complex hyperparameter tuning, so we used the default parameters without additional adjustments.

Table. Comparison of results between CatBoost and logistic regression

| **Logistic regression** |  | **CatBoost** |  |
| --- | --- | --- | --- |
| Top 5 Predictors | Coefficient, CI | Top 5 Predictors | SHAP value |
| Born in World War I | -20.053, [-4714.747, 4874.640] | Health before 15 | 0.264 |
| Cold running water | -0.355, [-0.413, -0.298] | Cold running water | 0.093 |
| Health before 15 | 0.268, [0.247, 0.290] | Relationship with father | 0.090 |
| Hot running water | -0.209, [-0.280, -0.138] | The number of rooms at 10 | 0.087 |
| Biological father lived | -0.153, [-0.230, -0.076] | Hot running water | 0.085 |

Notes: CI represents confidence interval. SHAP value measure the importance of each predictor variable in predicting the outcome, and a higher value indicates a greater importance of that predictor in the ML algorithm. For comparison with CatBoost, we employed logistic regression without any parameter tuning, treating it as a traditional model even though it is a ML algorithm.

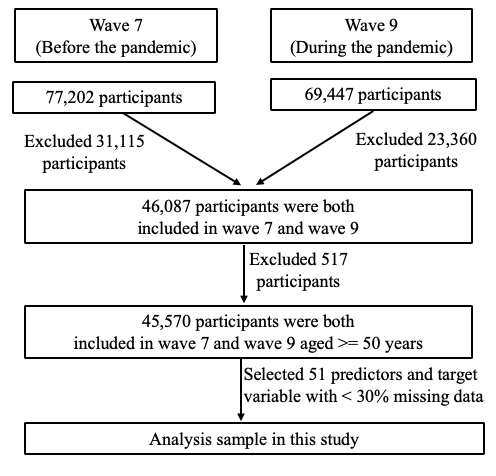

**Fig. Flow chart of the analytic sample**
